# The World According to Girls: A Qualitative Study of School, Work, and Identity Among Adolescent Girls and Young Women Living with HIV in Ghana

**DOI:** 10.1101/2025.07.22.25331754

**Authors:** Veena Bhagavathi, Charles Martyn-Dickens, Sheila Agyeiwaa Owusu, Gustav Nettey, Jessica Bedele, Ama D. Sarfo, Michelle Munyikwa, Cheryl A. Moyer, Anthony Enimil, Leah Ratner

## Abstract

**Background:** Adolescent girls and young women (AGYW) living with HIV in Ghana face multiple, intersecting forms of marginalization. Beyond the clinical management of HIV, little is known about how they construct meaning, navigate identity, and imagine their futures within structural contexts shaped by stigma, gender inequity, economic precarity and colonial legacies.

**Objective:** To explore how AGYW living with HIV in Ghana understand and negotiate their social identities in work and school. We then aimed to understand how their lived experiences at school and work are shaped by broader systems of power.

**Methods:** This qualitative study drew on semi-structured interviews with AGYW (ages 11–24, N = 24) receiving HIV care in Kumasi, Ghana. Data were coded both inductively and deductively. Themes were interpreted through the Ghanian context using intersectionality, Critical Disability Studies, spoiled identity theory and African Feminist decolonial theory. The analysis was conducted iteratively and reflexively, with attention to positionality, gender, and structural power dynamics.

**Results:** Seven major themes were identified: (1) social support; (2) concrete plans for the future; (3) unattainability of the future; (4) coping via detachment; (5) need for privacy and confidentiality; (6) role as arbiter of HIV information; and (7) financial stress. Across these themes, AGYW described dynamic processes of identity negotiation, moral and emotional labor, and structural constraint. HIV was rarely the sole barrier—rather, it intersected with gender norms, family dynamics, age hierarchies, economic marginalization, and misinformation to shape participants’ social worlds. Some participants coped through detachment or concealment, while others reclaimed agency through caregiving roles, education, or aspirational goals.

**Conclusions:** AGYW living with HIV in Ghana are not only navigating a chronic illness but also resisting a layered matrix of social and structural injustice. Their stories reveal both vulnerability and strategic agency. Interventions and policy must go beyond biomedical care to address stigma, provide confidential and affirming school and work environments, and offer structural supports for emotional, educational, and economic well-being.

**What is already known on this topic:**

- Adolescent girls and young women (AGYW) in sub-Saharan Africa represent a disproportionately high share of new HIV infections and face intersecting structural barriers related to gender, age, and stigma.
- Existing literature has shown that AGYW living with HIV often experience interruptions in education, social exclusion, and reduced employment opportunities.
- While some studies have explored stigma or psychosocial distress, few have examined how AGYW themselves construct meaning around their identities or futures.
- Intersectionality is frequently cited in global health but rarely operationalized in qualitative research in low- and middle-income countries (LMICs), and even less so alongside frameworks like Critical Disability Studies (CDS) or spoiled identity theory.

**What this study adds:**

- This study uses intersectionality, CDS, and spoiled identity theory in the context of Ghanaian culture to provide a nuanced analysis of how AGYW living with HIV experience and seek to overcome layered stigma in domains such as education, employment, romantic relationships, and family life.
- It reveals that HIV is not only a biomedical diagnosis, but a social identity entangled with broader systems of age-based power, ableism, gendered expectations and colonial identities.
- AGYW demonstrated agency through emotional detachment, strategic disclosure, and aspirational re-framing of their lives—navigating structural challenges with dignity, but often at high emotional cost.
- The findings show that resilience is often misinterpreted as individual strength when, in fact, it is evidence of institutional failure to provide supportive and affirming systems.

**How this study might affect research, practice or policy:**

- This study highlights the need for gender-and disability-responsive HIV programming that extends beyond adherence and viral suppression to address the moral, emotional, and social dimensions of living with HIV.
- It offers a model for applying intersectionality and CDS in qualitative research in LMIC settings, providing a replicable analytic approach for future studies.
- It calls on health and education systems—including institutions like the Ghana Education Service—to design inclusive trauma-informed, and decolonial policies that affirm the full humanity and aspirations of AGYW living with HIV.
- By recognizing AGYW as moral actors and arbiters of knowledge, the study suggests that program and policy design should be co-created with AGYW, not merely for them.

## INTRODUCTION

Over 80% of adolescents living with HIV reside in sub-Saharan Africa, amounting to an estimated 1.4 million young people.^1–3^ Among them, adolescent girls and young women (AGYW) represent the fastest-growing subgroup, comprising just 10% of the global population but accounting for 25% of new HIV infections in 2017.^3^ Gender disparities become particularly stark during adolescence: AGYW in sub-Saharan Africa are infected at a rate six times that of their male peers.^4^ While AIDS remains one of the leading causes of adolescent mortality in the region, the increased availability of antiretroviral therapy (ART) has shifted HIV from a terminal illness to a manageable, chronic condition.^5^

However, for AGYW, living with HIV is never merely a matter of managing a viral infection—it is also a profound social experience, marked by overlapping and often invisible forms of vulnerability. Studies show that AGYW living with or even exposed to HIV face lower educational attainment, greater school disruption, and higher levels of social exclusion^6,7^. At the same time, schools remain crucial sites for identity development, psychosocial support, and fostering a sense of future orientation. Disclosure within the school setting, though fraught with risk, can also open the door to belonging, trust, and connection.

Educational access and attainment are closely linked to long-term health and economic outcomes; for AGYW, they are also pathways to autonomy, dignity, and hope.^8^

AGYW living with HIV occupy a unique intersection of structural inequities—negotiating the challenges of adolescence, the constraints of gendered expectations, and the stigma of a chronic illness. While there has been some research examining the correlations between HIV and school or employment outcomes^9-10^, little is known about how AGYW themselves interpret, navigate, and make meaning of their diagnoses in these settings. Thus, we wanted to better understand, what does it mean to grow up with HIV in Ghana in school and at work? How do AGYW imagine their futures in terms of employment or education, respond to stigma in those settings, and reconcile aspirations in these contexts, within known social constraints?

To explore these questions, we employed an analytical approach grounded in intersectionality, a framework developed through Black feminist thought and coined by Kimberlé Crenshaw.^11^ Intersectionality challenges single-axis explanations of identity by recognizing that aspects such as gender, age and ability, interact to produce distinct lived experiences. Though increasingly cited in public health discourse^12^, intersectionality remains underutilized in both research design and policy implementation in low- and middle-income country (LMIC) settings, including much of sub-Saharan Africa^13^. As noted by scholars^14,15^, intersectionality in African contexts must also contend with the legacies of coloniality, structural adjustment, and donor-driven health agendas that often fail to reflect local realities and priorities and thus was interpreted through the Ghanian context.

While intersectionality initially guided the study design and informed the development of the interview guide, the complexity and depth of the data necessitated an expanded analytical framework. As analysis progressed, we incorporated Critical Disability Studies (CDS)^14^-^15^ to further interrogate the experiences of AGYW living with HIV in Ghana. CDS challenges deficit-oriented and medicalized models of disability and instead emphasizes sociopolitical understandings of impairment and marginalization. Although HIV is not always classified as a disability, especially in Ghana, we sought to critically reframe it as such to underscore the importance of this identity within broader equity and rights-based discourses. According to the Americans with Disabilities Act (ADA)^16^ and the Centers for Disease Control and Prevention (CDC)^17^, disability is defined as ‘a physical or mental impairment that substantially limits one or more major life activities’. AGYW living with HIV often meet these criteria, particularly as they navigate the compounded challenges of dynamic and/or invisible disabilities. Invisible disabilities refer to physical or mental health conditions that are not immediately apparent to others^18^, while dynamic disabilities are characterized by variability in symptom severity over time^19^. As Swoniska et al. note, individuals with invisible disabilities often face heightened challenges in accessing support due to nondisclosure and the lack of visible indicators of their condition^20^. Given the fluctuating nature of HIV and its general invisibility, we found that CDS offered a critical lens through which to understand how AGYW in Ghana experience and embody intersecting forms of marginalization.

CDS highlights the societal, cultural, and infrastructural systems that render individuals ‘disabled.’^14,15^ In Ghana and many other LMIC contexts, chronic health conditions (and invisible disabilities) such as HIV are rarely conceptualized as requiring structural accommodation^21^. Instead, they are often framed as personal moral failures, fueling internalized shame and limiting opportunities for support and understanding. Stigma theory^22^, coined by Goffman in 1963, examines the concept of ‘spoiled identity’ defined as the negative consequences of shame that emerge from that identity, specifically the concepts of enacted, anticipated, and internalized stigma. We used these definitions to guide further our understanding of how AGYW manage their identities through silence, disclosure, resistance, and adaptation.

Because we began with an intersectional lens, we noticed that many participants described feeling blocked from opportunities—what could be seen as a kind of ‘glass ceiling.’ These limits often seemed tied to internalized colonial ideas about what girls and women can or should become, we drew on African feminist and decolonial frameworks^23–25^, which challenge Western ideals of autonomy, productivity, and femininity, and call attention to the ways that colonial legacies continue to shape health policy, education, and even AGYW’s own aspirations. In contexts where silence is a form of survival, AGYW living with HIV must navigate expectations of secrecy, moral worth, and responsibility — expectations often shaped by institutions that fail to recognize them as full social actors.

This study aimed specifically to examine the lived experiences within the educational and economic contexts of this population, with the ultimate goal of advocating for inclusive policies. Through this work, we aimed to examine HIV not simply as a health condition, but as a social identity enmeshed with gender, class, age, ability and colonial histories. By focusing on the voices and agency of AGYW themselves, we hope this informs ability and gender-responsive approaches to education, economic attainment and social policy—particularly for institutions like the Ghana Education Service and the Ministries of Education and Health. Rather than framing these AGYW as passive players in their educational and occupational futures, our study positions them as captains of their own lives: navigating silence and surveillance, reimagining their worth, and crafting possibilities amidst constraints.

## METHODS

### Study Site

This study was conducted at the Komfo Anokye Teaching Hospital (KATH), a tertiary referral centre located in Kumasi, Ghana. KATH is the second-largest hospital in the country and serves as a major referral site for the middle belt and northern regions. Participants were recruited from the Child and Adolescent HIV Clinic, which provides longitudinal care for about 900 individuals living with HIV from infancy through young adulthood. At the time of study, the clinic was the only specialized facility of its kind in Ghana, drawing patients from approximately a third of the country.

### Study Population and Recruitment

Eligible participants included AGYW living with HIV, all with vertical acquisition, between the ages of 12 and 24 years who were aware of their HIV diagnosis and able to communicate in English or Twi.

Individuals were excluded if they were unaware of their diagnosis, unable to provide consent or assent, or unwilling to participate in the interview process.

Recruitment and interviews took place, in person, over a 12-month period from March 2024 to February 2025. Using purposive sampling, we aimed to capture a range of experiences across age groups and educational or employment contexts. Based on data suggesting saturation is achieved in 9-17 interviews^26^, we initially targeted 24 participants across two stratified age groups (10-19 and 20-24). Data collection continued until thematic saturation was reached in both groups —that is, the point at which no new themes, patterns, or insights were emerging from subsequent interviews.

Participants were recruited confidentially by trained staff at the HIV clinic. Informed consent or assent was obtained in accordance with ethical guidelines. Participants were informed that their participation was voluntary, confidential, and would not affect their care.

### Ethical Approval

Ethical approval for this study was obtained from the Komfo Anokye Teaching Hospital Institutional Review Board prior to the initiation of research activities. (KATH IRB/AP/040/24) The KATH IRB served as the IRB of record for this research.

### Study Design and Data Collection

This was a qualitative study using semi-structured interviews (N=24). Though a total of 24 interviews were conducted, four initially stated that they were unaware of their diagnosis and were excluded – however after discussion with their primary provider, it was confirmed that they did but were just uncomfortable sharing that with strangers—and their interviews were re-included and analyzed. No participants declined to participate and no one was present other than interviewers and participants.

Interviews were conducted in person by trained research assistants who were fluent in both English and Twi. To minimize participant burden, interviews were scheduled during routine clinic visits and conducted in a private area of the clinic. The interview guide was piloted prior to beginning. Each interview lasted approximately 15–30 minutes and was conducted in the participant’s preferred language (English, Twi, or both). No one besides the participant and interviewer was present during interviews and there were no repeat interviews. Interviews followed a semi-structured guide developed by the research team, incorporating open-ended questions and targeted probes related to education, employment, HIV-related challenges, and coping strategies. Interviews were audio-recorded with permission, and then transcribed to English, de-identified and anonymized prior to analysis. Field notes were also taken and uploaded to the coding software for analysis.

### Data Analysis

Audio recordings were translated and transcribed into English by trained bilingual team members familiar with the clinical and cultural context. Transcripts were analyzed using thematic analysis. No transcripts were returned for correction. Data was initially stratified by two age groups (10-19 and 20-24), however there was significant intersection across both, and were merged. Coding was conducted in Taguette software^27^ using a hybrid inductive and deductive approach. Two independent researchers coded the transcripts (LR,VB), then met to resolve discrepancies and finalize a shared codebook. A third reviewer assessed the codebook for contextual accuracy and cultural relevance (SAO). Codes were then organized into themes and subthemes. Thematic was reached when no new codes emerged from subsequent transcripts. Saturation was assessed iteratively during data collection and analysis and was jointly agreed upon by members of the research team (LR, SAO, VB). Thematic saturation was reached after 16 interviews. Saturation was assessed inductively and in line with the study’s epistemological approach, recognizing that in interpretivist frameworks, saturation reflects sufficient depth and diversity of meaning rather than exhaustive category listing. This manuscript was developed in accordance with the Consolidated Criteria for Reporting Qualitative Research (COREQ) checklist to ensure transparency and rigor in qualitative reporting.

### Reflexivity

The research team engaged in regular and intentional discussions throughout the study to identify and reflect on personal and collective blind spots related to gender, ableism, colonialism, and sexism. Those who conducted interviews (VB and JB) are both female-identifying resident physicians and had no prior relationship with the participants. They both have introductory knowledge of qualitative research. Coders (LR, SAO) have terminal degrees and have more advanced training in qualitative research.

All authors had some lived experience with sexism, colonialism, ageism and ableism—those with explicit experience led discussions and analyses around those experiences. We engaged in intentional, iterative conversations to examine how our positionalities might shape our assumptions and interpretations. Team members with more experience in the Ghanaian context provided critical cultural and linguistic guidance. In contrast, those with lived experience of chronic illness contributed insight into the affective and embodied aspects of coping with long-term health conditions. Decisions regarding study leadership, authorship, and analysis were made with consideration of both identity and power dynamics. By acknowledging and embracing our different perspectives, the team attempted to address potential blind spots, challenge assumptions, and co-construct meaning in a way that reflected both local context and the broader systemic forces shaping the lives of participants. As a research team, we understand that no single identity defines a person, and that the way different identities intersect can create unique experiences of marginalization—some of which we may still overlook.

## RESULTS

N=24 interviews were completed and analyzed. The ages of respondents ranged from 11-24, with a mean age of 16.8 years old and mode age of 15 years old. 14 participanrs (58%) were less than 19 years old, and N=10 (42%) were 20 years old or greater. Thematic analysis of interviews with AGYW living with HIV in Ghana revealed a dynamic interplay of social, emotional, and structural forces shaping their lives. Participants exhibited resilience, purpose, and agency, while also navigating stigma, financial hardship, and gendered expectations. Seven themes emerged: (1) social support, (2) concrete plans for the future, (3) unattainability of the future, (4) coping via detachment, (5) need for privacy and confidentiality, (6) arbiter of information and (7) financial stress.

**Table 1:**
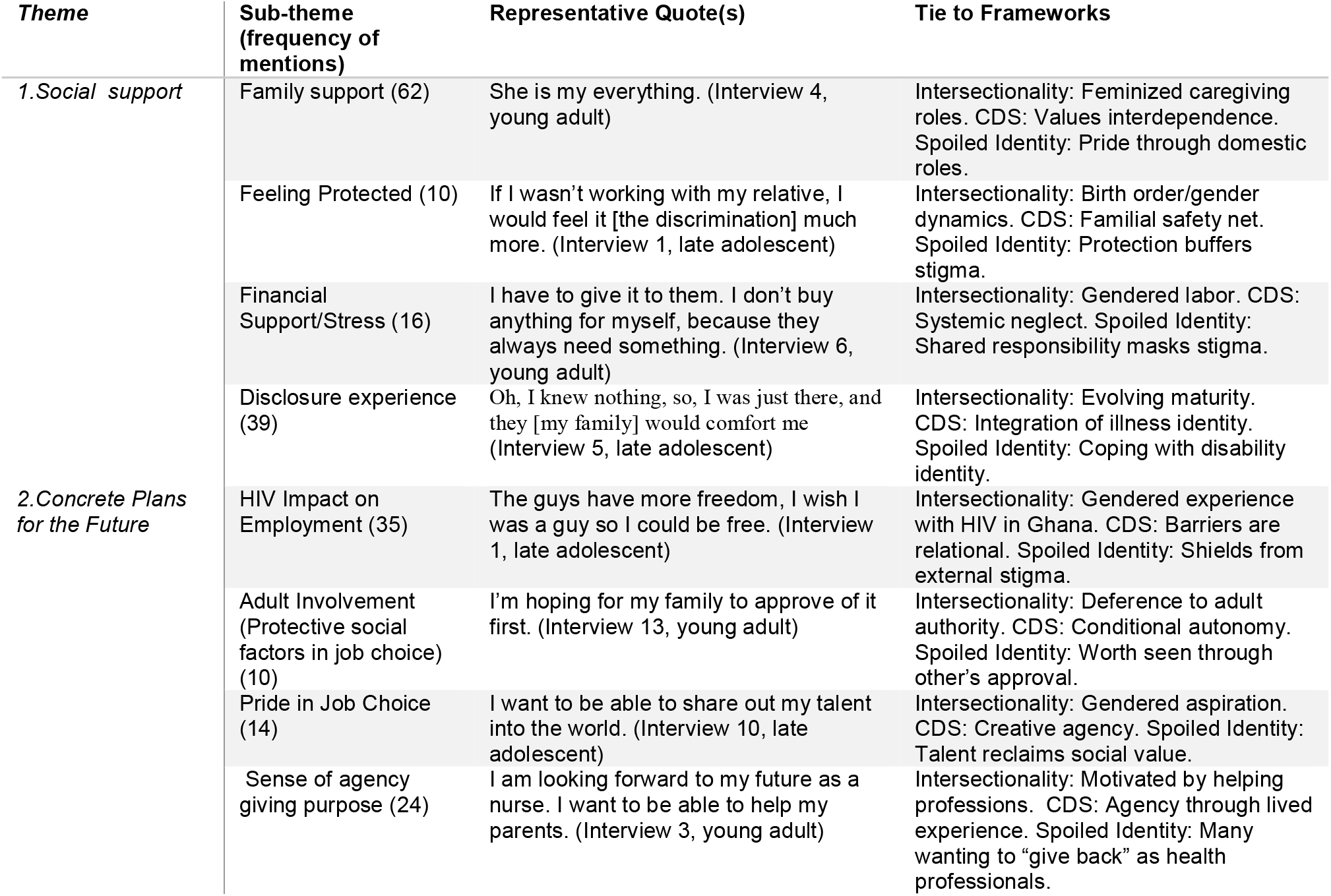
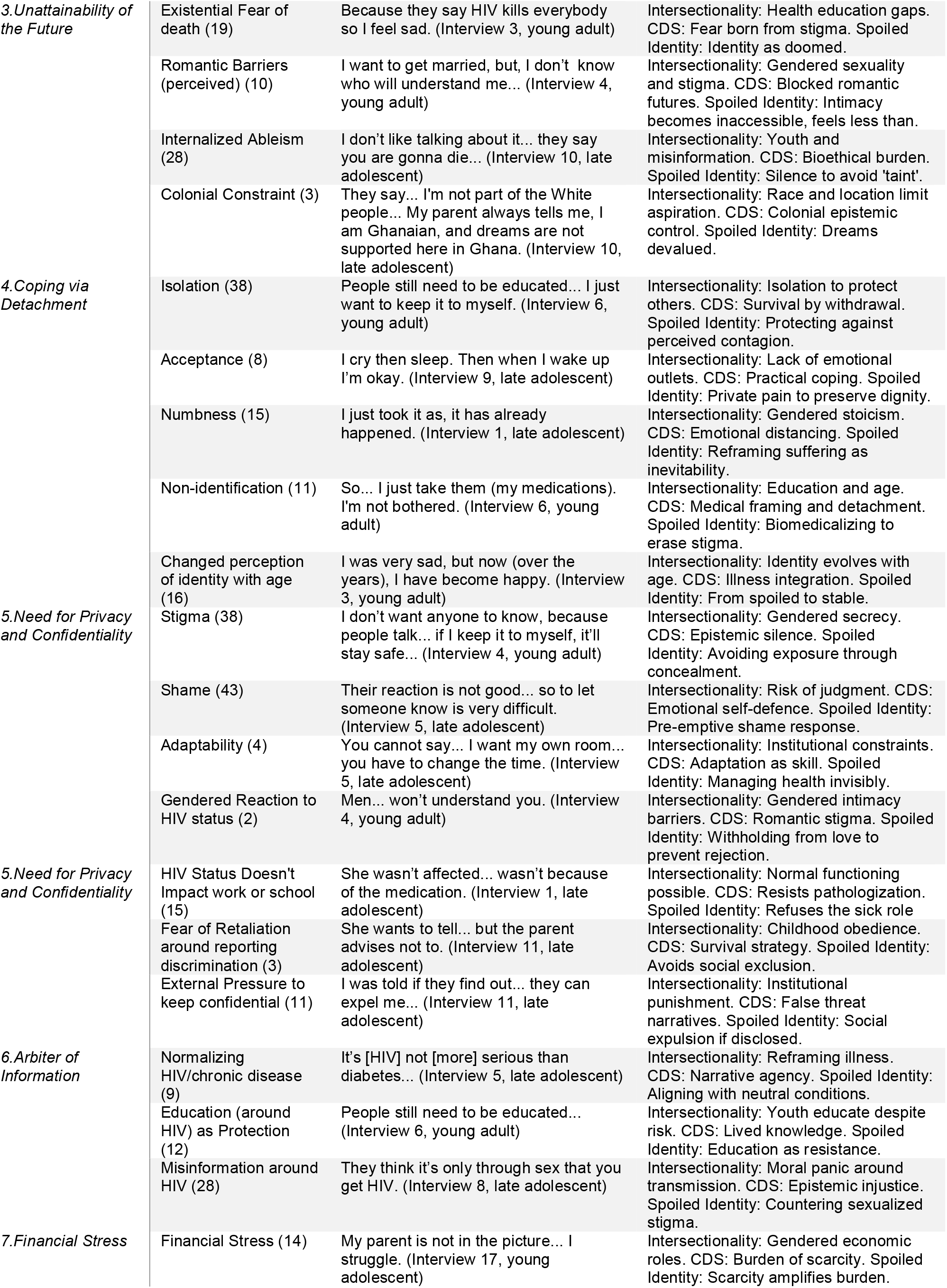
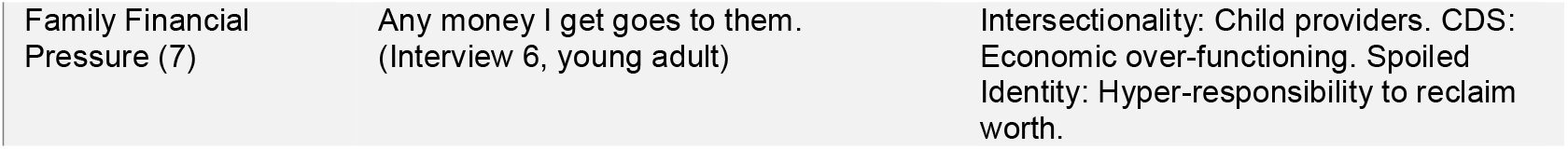
Results -- Themes, Quotes and Ties to Frameworks.

### Social Support

Though social support was a major theme, the majority of social support was drawn from family, with 62 mentions of family support. Family was central in helping participants navigate their diagnosis as many drew strength from caregiving roles, describing being proud or happy that they were able to take care of their siblings or their families. Ten mentioned feeling protected by their social support, and often specifically by their family. Much of the conversation illustrating this was centred around the AGYW’s disclosure experience (which was mentioned 39 times) and how their families would comfort them.

Interview 6, a young adult, who is currently working, described the experience around disclosure and how her family comforted her *‘She said, if you take the drugs, you will not see the virus again. So I thought, It’s an easy thing, so I’ll just take them*.*’*

### Concrete Plans for the Future

Participants articulated both aspiration and discouragement. There were 35 mentions of HIV having some impact on work or school. Some talked about losing motivation upon witnessing family members have to stop schooling or working. However, similar to the previous theme, there were 10 mentions of a family member having a protective factor in their future job choice, for example, choosing to work with relatives, so they wouldn’t have to disclose their status to their employer. However, there was also a lot of excitement and pride about the future and their choices. There were 14 times when pride was noted when describing their job choice for the future, and 24 times that there was some sense of agency in their school or work choice that seemed to give purpose. Most noted that they wanted to be a doctor or a nurse, with many citing inspirations from their relationships with the current HIV doctors and nurses.

### Unattainability of the Future

Though there was obvious pride in feeling able to make decisions that shape one’s future, there was also a duality that AGYW felt parts of that future were unattainable due to their intersecting identities. Decisions around the type of work they hoped to do in the future, were often shaped by anticipated stigma (internalized ableism (28 mentions) and colonial constraint (3 mentions)). The internalized ableism was often illustrated by the AGYW sharing that they were not worthy of being at school or at work due to their condition. Often citing that they didn’t want to be the subject of pity or othering. This was often heightened when it intersected with colonial constraint, often describing unattainable futures not only because of their disease but because of the constraints of Ghana. Interview 10 (late adolescent) demonstrated this well when she said, *‘For me, I don’t think that’s possible. I don’t know. They say that my dream, I’m not part of the White people, but that I’m here in Ghana. In Ghana, such dreams aren’t supported here, so I’d better find another dream. My parent always tells me that*.*’*

Some participants feared their health would deteriorate before they could realize their goals. Others struggled to articulate any vision of the future because of this fear. Narratives were shaped by messages from schools and peers. One participant recalled being told her chronically ill friend was told that she would die, so there is no need to go to school. As part of what felt unattainable, was being in a romantic relationship, which was mentioned 10 times. Interview 4, a young adult said, Yeah…*I just dropped my guy, my ex. I asked him [hypothetically], if you found out that the girl you are in love with is having HIV…–he said no no no I will never [date someone with HIV]. I felt so bad*.*’*

### Coping via Detachment

Detachment emerged as a coping strategy, and more specifically themes around isolation (mentioned 38 times), acceptance (mentioned 8 times), numbness (15 times) and participants saying they didn’t identify with the disease state (11 times)—all as examples of how they used detachment as a coping strategy. One participant stated, *‘I just want to take my medications, so I don’t affect anyone*.*’* (Interview 6, late adolescent). Others turned inward: *‘Cry then sleep. Then when I wake up, I’m okay*.*’* (Interview 9, young adult). Given the developmental need at this age to be social, especially with peers, it was notable that most participants noted at least once that they don’t feel comfortable with even their best friends knowing their status and then further reflected on how that has influenced intense isolationism. This felt like a interesting juxtaposition, considering the communal nature of Ashanti culture.

### Need for Privacy and Confidentiality

Stigma was a theme found in most interviews with 38 mentions. Secrecy was a key strategy to manage stigma. Often further describing the reasons behind their need to isolate, coupled with internalized ableism, participants described wanting to keep away from others, both for self-preservation but also because of a belief that that was also better for their peers. Fear of being perceived as contagious drove also concealment, and parents also often encouraged isolation and non-disclosure at school. This not surprisingly, also brought up a lot of shame in their identities and perceptions of who they are in this world. Shame was mentioned 43 times. Two people also talked about the need for confidentiality that seemed especially tied to their gender, one noted that she was intentionally keeping it from her parent which she implied she needed to do because they would overreact. Another AGYW said her need for confidentiality was because she knew she needed to get married one day, and she was worried this would prevent that from happening, but also implied this might be a double standard had she been born male.

However, there were also some really positive adaptations were made. Four noted how they adaptation their day at school or work in a way that allows for confidentiality in taking medications but causes less stress or isolation. Because of many of these adaptations, 15 noted that their HIV status doesn’t actually impact school or work.

However, retaliation and pressure (from external sources) to keep their status and illness confidential while at work or school were also noted. Three people mentioned explicit fears of retaliation if their teachers found out. Interview 11, a late adolescent, noted a classmate getting transferred for a similar disclosure, and therefore was very scared about being found out. However, Interview 13, a young adult, who is currently working said, ‘*At first, when you are having the HIV, they fear for you to work with them because they think that maybe if it’s a little something cut, or you got a cut. …But for now [its better] everybody works*.*’* Showing some evolution in what she perceived vs what was actually happening.

### Arbiter of Information about HIV

Despite stigma, many participants assumed roles as informal educators, often as an attempt to both protect themselves as well as because others assumed them to be expert on the experience of being an AGYW with HIV. Nine noted times when they attempted to normalize HIV and chronic disease. *‘It’s not all that serious than diabetes and cancers*… *it can at least control it*,’ (Interview 5, young adult). This often parlayed into HIV education that they provided to their peers (noted 12 times). However, it was mentioned 28 times that there is misinformation about HIV being spread in their work or school, often adding to shame or stigma.

### Financial Stress

Poverty shaped daily decisions and long-term goals. There were 14 mentions of financial stress impacting their identity at work or school. Some prioritized family over themselves: ‘*I don’t buy anything for myself because they always need something*.*’* (Interview 6, late adolescent). There were 7 mentions of giving money to family members before taking care of themselves (and their HIV-related needs). Some coped by assuming hyper-responsibility as a form of social redemption, but were often unsupported in rest or self-care.

### Collective Reflection

Across all themes, participants navigated tension between managing stigma and asserting agency. Their narratives reflected identities in motion, reworked through context, silence, and emotion. HIV emerged as a deeply social experience, negotiated daily with extraordinary strength.

## DISCUSSION

This study aimed to explore how AGYW living with HIV in Ghana experience school and work within systems shaped by stigma, inequality, and structural exclusion. By using intersectionality, CDS, and African feminist theory, we illuminate how HIV is not just a medical condition but a deeply social identity—one that interacts with age, gender, and poverty to shape AGYW’s futures. In addition, this analysis was guided by the Ghanian context and the lived experience of living with chronic illness. These findings contribute to a growing body of literature that challenges dominant biomedical and individualistic framings of HIV in adolescence.

### Intersecting Marginalizations in Work and School

This study offers a nuanced look into how AGYW in Ghana living with HIV experience, negotiate, and overcome layered forms of marginalization—particularly in work and school settings. Our findings reveal that these experiences are shaped not only by HIV stigma, but by broader systems of power, including patriarchy, ableism, age-based hierarchy, and structural poverty. This aligns with prior work by Nutor et al^28^. and Asiedu et al,^29^ which show how stigma and shame intersect in the Ghanian context, with gender norms and family expectations, limiting educational and occupational opportunities. Intersectionality illuminated how overlapping identities—including adolescence, gender, HIV status, and poverty—created compounded barriers. In this way, HIV status functions as both a lived experience and a social marker, shaping what participants felt they could say, do, or become.

### Invisible Disability and Spoiled Identity in School and Work

Though a few participants described physical disability as a barrier to work or school, the more common narrative was of HIV as an invisible and dynamic disability. As CDS scholars note, the invisibility of certain chronic conditions intensifies stigma by forcing concealment, limiting access to support, and increasing internalized shame.

Participants repeatedly described exhaustion and internalized stigma. These examples demonstrate that even without visible symptoms, AGYW must navigate their condition carefully, constantly monitoring when, where, and how to disclose—or hide—their status. This was further contextualized by a previous paper written by several of this paper’s authors, that though internalized stigma is common this population, the collective nature of Ashanti culture can be an effective antidote.^30^

These experiences also translated into fears about future relationships. These accounts mirror Shamrock & Ginn’s articulation of ‘sexual ableism,’^31^ where disabled youth are rendered invisible in discourses of romance, family, and intimacy. Participants internalized this narrative, often seeing themselves as unworthy or undesirable partners due to HIV, a notion reinforced by discriminatory attitudes from others. Ghanaian culture puts high emphasis on marriage and family, which can be ‘othering’ to those who believe that is an attainable goal.

School was often framed as a site of mixed support and harm. Misinformation about HIV persisted, contributing to exclusion and eroding future aspirations. Messages received from school and peers often echoed the idea that chronic illness rendered one’s future void. These internalized narratives contributed to what Goffman called ‘spoiled identity’^22^—a sense that one’s future is disqualified by moral or physical inadequacy.

### Reframing Illness and Reclaiming Agency

Despite these constraints, several AGYW reframed their experience of illness into one of meaning, motivation, and purpose. Career aspirations were often inspired by their exposure to healthcare through the HIV clinic.

These aspirations echo findings by Burke et al., who found that participation in HIV programs cultivated youth leadership and vocational agency,^32^ which was further hypothesized in a previous mixed-methods study by several of this paper’s authors, in the same clinical and regional context.^30^ Within CDS, these moments are understood as reclaiming spaces historically marked by stigma. Rather than retreating from medical spaces that signified marginalization, AGYW reimagined these institutions as platforms for empowerment.

Participants also derived identity from gendered caregiving roles. This framing echoes African feminist theory (e.g., Tamale^33^), which challenges American feminist notions of empowerment --- by household work and caregiving as a form of agency and relational power. Rather than viewing caregiving as constraint, these AGYW viewed their support roles as valuable, moral contributions to family and community.

### Strategic Adaptation and the Emotional Cost of Resilience

AGYW engaged in significant strategic adaptation to navigate systems not designed to accommodate their health, emotional, or educational needs. Participants described covert medication routines emotional suppression and hyper-responsibility.

These survival strategies demonstrate not only resilience but the cost of living without institutional support. This is a core tenet of CDS--resistance to celebrating individual adaptation while ignoring the injustices that create the need to adapt in the first place. CDS reframes these behaviors not as exceptionalism^34^, but as the inevitable result of unjust systems—those that do not provide trauma-informed care, confidentiality in school settings, or mental health support. Spoiled identity theory^22^ helps us see how AGYW worked to ‘repair’ their identities in a society that routinely questioned their moral worth. Yet this repair often required emotional sacrifice, silence, and the suppression of their own needs.

Still, some AGYW refused these constraints. Their resistance, whether through education, ambition, or pride in caregiving, echoes African feminist calls for epistemic refusal^33^—the rejection of externally imposed narratives and the assertion of one’s own meaning. These AGYW were not passive recipients of care or stigma; they were moral agents shaping, resisting, and sometimes transcending the systems around them.

## Data Availability

All data produced in the present work are contained in the manuscript

## Conclusion: Implications for Health Justice

This study contributes to the growing literature that conceptualizes HIV not solely as a disease, but as a complex, socially-situated identity—especially for AGYW navigating work and school. Intersectionality, CDS, spoiled identity theory, and African feminist frameworks reveal how these young women are multiply burdened and multiply resilient. HIV in this context is both a health condition and a lens through which gender, class, age, and colonial legacies interact to shape experience.

Our findings underscore the need for health and education systems in Ghana to create disability- and gender-responsive services that build off the healing power of the Ghanaian emphasis on community. These include confidential school-based accommodations, safe spaces for emotional processing, economic support for caregiving AGYW, and health messaging that does not frame HIV as a moral failure. This would allow them to once again have the freedom to make career and education choices based on factors other than their marginalized identities. AGYW must be viewed not only as recipients of care but as agents of knowledge, caretakers, educators, and aspirational thinkers.

A just world would not simply ask them to cope; it would rise to meet their needs. Their identities should not be managed around systems—they should help shape and redesign those systems. HIV programming must be reimagined in ways that are participatory, intersectional, and responsive to the lived realities of AGYW. By centering their experiences, we can move closer to a world where care, dignity, and opportunity are accessible to all.

## References

1. Magobo RE, Mabaso M, Jooste S, et al. 95-95-95 HIV indicators among children younger than 15 years in South Africa: results from the 2017 national HIV prevalence, incidence, behaviour, and communication survey. AIDS Res Ther. 2025;22(1):6. doi:10.1186/s12981-024-00691-8

2. HIV.Gov. Global statistics. Updated 7 Feb 2025. https://www.HIV.gov.

3. Sam-Agudu NA, Folayan MO, Ezeanolue EE. Seeking wider access to HIV testing for adolescents in sub-Saharan Africa. Pediatr Res. 2016;79(6):838–845. doi:10.1038/pr.2016.28

4. Dellar RC, Dlamini S, Karim QA. Adolescent girls and young women: key populations for HIV epidemic control. J Int AIDS Soc. 2015;18(2 Suppl 1):19408. doi:10.7448/IAS.18.2.19408

5. Oguntibeju OO. Quality of life of people living with HIV and AIDS and antiretroviral therapy. HIVAIDS Auckl NZ. 2012;4:117–124. doi:10.2147/HIV.S32321

6. Ngwenya N, Smith T, Shahmanesh M, et al. Social Categorisation and Social Identification: The Mediating Role of Social Isolation and Loneliness in Adolescents Living with HIV. Int J Behav Med. 2024;31(3):459–467. doi:10.1007/s12529-023-10205-x

7. Mhungu A, Sixsmith J, Burnett E. Adolescent Girls and Young Women’s Experiences of Living with HIV in the Context of Patriarchal Culture in Sub-Saharan Africa: A Scoping Review. AIDS Behav. 2023;27(5):1365–1379. doi:10.1007/s10461-022-03872-6

8. Mulwa S, Chimoyi L, Agbla S, et al. Impact of the DREAMS interventions on educational attainment among adolescent girls and young women: Causal analysis of a prospective cohort in urban Kenya. Ortega JA, ed. PLOS ONE. 2021;16(8):e0255165. doi:10.1371/journal.pone.0255165

9. Maulsby CH, Ratnayake A, Hesson D, Mugavero MJ, Latkin CA. A Scoping Review of Employment and HIV. AIDS Behav. 2020;24(10):2942–2955. doi:10.1007/s10461-020-02845-x

10. Yu Y, Chen Z, Huang S, Chen Z, Zhang K. What determines employment quality among people living with HIV: An empirical study in China. PloS One. 2020;15(12):e0243069. doi:10.1371/journal.pone.0243069

11. Crenshaw, Kimberle (1989) ‘Demarginalizing the Intersection of Race and Sex: A Black Feminist Critique of Antidiscrimination Doctrine, Feminist Theory and Antiracist Politics,’ University of Chicago Legal Forum: Vol. 1989, Article 8. Available at: https://chicagounbound.uchicago.edu/uclf/vol1989/iss1/8.

12. Bauer GR. Incorporating intersectionality theory into population health research methodology: Challenges and the potential to advance health equity. Soc Sci Med. 2014;110:10–17. doi:10.1016/j.socscimed.2014.03.022

13. Bauer GR, Churchill SM, Mahendran M, Walwyn C, Lizotte D, Villa-Rueda AA. Intersectionality in quantitative research: A systematic review of its emergence and applications of theory and methods. SSM - Popul Health. 2021;14:100798. doi:10.1016/j.ssmph.2021.100798

14. Vehmas S, Watson N. Moral wrongs, disadvantages, and disability: a critique of critical disability studies. Disabil Soc. 2014;29(4):638–650. doi:10.1080/09687599.2013.831751

15. Shildrick, M. 2012. ‘Critical Disability Studies: Rethinking the Conventions for the Age of Postmodernity.’ In Routledge Handbook of Disability Studies, Edited by N. Watson, A. Roulstone and C. Thomas, 30–41. London: Routledge.

16. Americans with Disabilities Act of 1990, 42 U.S.C. § 12101 et seq. (1990).

17. Chronic Disease Definition: US Department of Health and Human Services. Multiple Chronic Conditions: A Strategic Framework; 2010.

18. Matthews, C. K., & Harrington, N. G. (2000). Invisible disability. In Braithwaite D. O. & Thompson T.L. (Eds.), Handbook of communication and people with disabilities: Research and application (pp. 405–421). Lawrence Erlbaum.

19. Jakimovski D, Weinstock-Guttman B, Burnham A, et al. Dynamic disability measures decrease the clinico-radiological gap in people with severely affected multiple sclerosis. Mult Scler Relat Disord. 2024;87:105630. doi:10.1016/j.msard.2024.105630

20. Sowińska A, Pezoa Tudela R. Living with invisible medical disabilities: experiences and challenges of Chilean university students disclosed in medical consultations. Int J Qual Stud Health Well-being. 2023 Dec;18(1):2221905. doi: 10.1080/17482631.2023.2221905. PMID: 37300841; PMCID: PMC10259308.

21. Gyimah EM, Dassah E, Opoku MP, et al. From legislation to actual health service: evaluation of health provisions in the disability law of Ghana by adolescents with mobility and visual impairments and their families. BMC Health Serv Res. 2024;24(1):1314. doi:10.1186/s12913-024-11611-x

22. Goffman, E. (1963). Stigma: Notes on the Management of Spoiled Identity. Englewood Cliffs, NJ: Prentice-Hall.

23. Nkenkana, Akhona. ‘No African Futures without the Liberation of Women: A Decolonial Feminist Perspective.’ Africa Development 40.3 (2015): 41–57.

24. Byrne, Deirdre. ‘Decolonial African Feminism for White Allies.’ Journal of International Women’s Studies 21.7 (2020): 37–46.

25. Okech, Awino. ‘African Feminist Epistemic Communities and Decoloniality.’ Critical African Studies 12.3 (2020): 313–329.

26. Hennink M, Kaiser BN. Sample sizes for saturation in qualitative research: A systematic review of empirical tests. Soc Sci Med. 2022;292:114523. doi:10.1016/j.socscimed.2021.114523

27. Taguette Software. https://www.Taguette.Org/about.html.

28. Nutor JJ, Gyamerah AO, Duah HO, et al. The association of HIV-related stigma and psychosocial factors and HIV treatment outcomes among people living with HIV in the Volta region of Ghana: A mixed-methods study. Sundararajan R, ed. PLOS Glob Public Health. 2024;4(2):e0002994. doi:10.1371/journal.pgph.0002994

29. Asiedu GB, Myers-Bowman KS. Gender Differences in the Experiences of HIV/AIDS-Related Stigma: A Qualitative Study in Ghana. Health Care Women Int. 2014;35(7-9):703–727. doi:10.1080/07399332.2014.895367

30. Omuojine JP, Martyn-Dickens C, Owusu SA, et al. Understanding depression, anxiety and stress in young people living with HIV in Ghana. Afr J AIDS Res. Published online October 21, 2024:1–9. doi:10.2989/16085906.2024.2370792

31. Shamrock OW, Ginn HG. Disability and Sexuality: Toward a Focus on Sexuality Education in Ghana. Sex Disabil. 2021;39(4):629–645. doi:10.1007/s11195-021-09699-8

32. Burke VM, Frimpong C, Miti S, et al. ‘It must start with me, so it started with me’: A qualitative study of Project YES! youth peer mentor implementing experiences supporting adolescents and young adults living with HIV in Ndola, Zambia. Zanoni BC, ed. PLOS ONE. 2022;17(2):e0261948. doi:10.1371/journal.pone.0261948

33. Sylvia Tamale, Decolonization and Afro-Feminism, Ottawa, Daraja Press, 2020, Pp. 420.

34. Hutcheon, E.DJ. & Wolbring, G. (2013). Deconstructing the resilience concept using an ableism lens: Implications for People with Diverse Abilities. Dilemata, 11, 235–252.

